# CFD-derived biomarkers in intermediate risk pulmonary embolism patients treated with mechanical thrombectomy

**DOI:** 10.64898/2026.07.09.26357404

**Authors:** M. Gilani, A. Barr, M. O. Al-Qadi, J. M. Szafron

## Abstract

**Background:** Acute pulmonary embolism (PE) is a leading cause of morbidity and mortality with persistent difficulties in choosing interventions and predicting outcomes for patients defined clinically as intermediate risk. Computational fluid dynamics (CFD) tools have been used to understand the hemodynamic environment and plan interventions in the pulmonary arteries across a variety of disease conditions. Several biomechanical metrics have been used to evaluate risk in narrowed vessels, including hemodynamic resistance, power dissipation, and fractional flow reserve (FFR). In this study, we evaluate differences in these CFD-derived biomarkers between healthy controls (HC) and intermediate risk, acute PE patients. Additionally, we examine the response of patient hemodynamics to mechanical thrombectomy and compare values of these biomarkers across post-intervention pressure status.

**Methods:** A CFD framework was developed to simulate patient-specific hemodynamics within the pulmonary vasculature identifiable from clinical imaging. The pipeline involved reconstructing three-dimensional (3D) structures of the pulmonary arteries and modeling blood flow with the finite element method. Patient-specific boundary conditions were derived from matching pre-intervention inlet mPAP to the patient’s measured value given their measured CO as steady inflow. Converged simulations allowed for precise quantification of primary hemodynamic characteristics (flow and pressure) as well as secondary flow phenomena, primarily wall shear stress (WSS) and simulated pressure metrics such as fractional flow reserve (FFR).

**Results:** Our simulations revealed significant elevations in resistance, power dissipation, and the number of vessels with low FFR in those patients with acute PE (n=6) compared to HC (n=3). Occlusions of hemodynamic significance were generally found in segmental pulmonary arteries. For patients with normalized pulmonary pressures post-thrombectomy (n=3), we found significantly higher proximal power dissipation and counts of low FFR vessels in comparison to those with elevated pressures after intervention (n=3). Distal resistance, which was derived from the portion of resistance attributed to the outflow boundary conditions, was significantly higher in patients with elevated pressures post-intervention. Across all PE patients, FFR count was significantly correlated with post-thrombectomy pulmonary pressure and cardiac index.

**Discussion:** CFD-derived biomarkers offer a promising tool for understanding disease severity in acute PE. Differences between HCs and acute PE patients reveal expected increases in metrics associated with proximal disease burden. Yet, in examining acute PE patients with varying post-intervention hemodynamics, we found that these metrics of proximal disease burden could also be useful to predict the efficacy of mechanical thrombectomy. Those patients with normalized pressures had higher values for proximal disease metrics and lower values for distal disease metrics than those with continued elevations in pressure. This suggests that accessibility of hemodynamically-significant emboli to thrombectomy may be useful as a predictor for outcomes.

## INTRODUCTION

Acute Pulmonary Embolism (PE) is one of the leading causes of cardiovascular related death, with 150,000-250,000 hospitalizations and 60,000-100,000 deaths per year in the United States [1]. A broad array of treatments are currently available, and decisions hinge heavily on clinical risk stratification. Currently, this stratification relies on clinical parameters independent of radiographic clot burden. AHA Class A and B (formerly low-risk) patients are those without evidence of hemodynamic compromise or cardiac dysfunction while class D and E patients (formerly high-risk) are those with incipient or frank cardiovascular failure. Both groups have well defined treatment pathways with the former relying on anticoagulant monotherapy and the latter proceeding to immediate systemic thrombolysis [2]. The management of Class C patients—those without hemodynamic compromise but with evidence of cardiac dysfunction—is partially dependent on a number of novel, minimally invasive treatment methods. Amongst these, mechanical thrombectomy (MT) shows promise as a safe and effective treatment paradigm. In STORM-PE, the only randomized clinical trial comparing MT to anticoagulation, MT demonstrated significantly greater reduction in RV/LV ratio at 48 hours when compared to anticoagulation alone, but did not detect a difference in mortality [3]. Despite these initial findings, STORM-PE data is inherently limited, and longitudinal data on the reduction in progression to CTEPD and Post-PE syndrome do not currently exist. Consequently, optimized patient selection may enhance the outcomes of these interventional therapies while limiting patient exposure to invasive and resource-intensive procedures. Current hypotheses regarding the pathogenesis of both CTEPD and Post-PE syndrome center around a combination of persistent proximal and microvascular obstruction, as well as small vessel arteriopathy. Residual obstruction has been demonstrated on SPECT imaging and correlates with clinical symptoms [4]. Small vessel arteriopathy is thought to be secondary to endothelial dysfunction as a result of nonphysiological wall shear stress from obstruction related aberrant flow [5]. However, identifying which patients are experiencing these critical hemodynamic derangements remains challenging, as current standard-of-care radiographic clot assessments provide purely anatomical visualizations and lack the physiologic data necessary to understand the true functional impact of the obstruction.

To bridge this gap between anatomical appearance and physiological impact, metrics such as Fractional Flow Reserve (FFR) and Cardiac Power Output (CPO) offer a compelling functional approach. In the management of coronary artery disease, FFR is widely established as a hemodynamic index that indicates physiological significance of a stenosis. Higher FFRs have been associated with less severe occlusions and better long term clinical outcomes [6]. Similarly, measures of CPO and stroke work are routinely utilized in heart failure and critical care settings to assess ventricular performance and the energetic toll of overcoming vascular resistance [7,8]. Applied to the pulmonary vasculature in the setting of acute PE, FFR can provide a physiologic, flow adjusted method of assessing hemodynamic disruption from individual embolic lesions. Similarly, calculating the vascular power dissipated across occlusions may offer a specific functional assessment of disease burden and its implications of cardiac function and right ventricular strain. Despite these benefits, a major barrier to their adoption in routine PE management is the difficulty of evaluating these functional markers non-invasively.

Computational fluid dynamics (CFD) modeling of the vasculature offers a potential solution to this problem. CFD can provide a noninvasive or minimally-invasive method for assessing hemodynamics by simulating fluid flow with physics-based numerical methods. By applying boundary conditions derived from measured quantities, such as resistance and inflow, hemodynamic values, including pressure and wall shear stress (WSS), can be simulated across a personalized, 3D domain of discrete finite elements for each patient [9,10]. Prior works have utilized CFD to simulate vascular biomechanics in disease conditions, such as predicting blood flow and FFR to better diagnose the severity of lesions in coronary artery disease [11,12]. CFD-based FFR has also been used as a minimally invasive method of predicting patient response to surgical intervention [13,14]. Similarly, assessment of different surgical geometries in congenital heart disease have allowed for analysis of vascular power dissipation, which impacts long-term cardiac function [15,16]. Recent progress has also been made in CFD modeling of the pulmonary circulation, such as in planning stent placement for peripheral pulmonary artery stenosis [17–19] However, there is still a need to evaluate their utility in understanding hemodynamic changes and predicting outcomes for patients with acute PE.

In this work, we used CT imaging and hemodynamic data from catheterization in patients with acute PE treated with MT to model the pulmonary vasculature within the open-source CFD program SimVascular. Through an iterative boundary condition tuning process, contributions to proximal vs distal resistance and power dissipation were identified. Additionally, secondary metrics were derived locally from the 3D simulation results in the proximal pulmonary arteries, including resistance, FFR, and power dissipation. Distinct differences were found between patients with post-procedural pulmonary hypertension (PPH) and those without (nPPH), which suggests a potential future role for CFD in predicting the response to MT for acute PE patients.

## METHODS

### Study Population

In accordance with a protocol approved by the University of Pittsburgh Medical Center (UPMC) Institutional Review Board, a retrospective study was conducted by selecting adult patients with intermediate-high risk PE [20], who underwent suction thrombectomy from a database of patients with diagnosed PE that presented to hospitals within the UPMC system between 2014-2025. Selected patients were required to have following set of pre-intervention right heart catheterization (RHC) data: heart rate, cardiac output (CO), and pulmonary artery pressures. Additionally, patients were required to have post-intervention pulmonary artery pressures to assess response to treatments. Patients were excluded if they demonstrated evidence of acute hemodynamic decompensation or possessed other underlying cardiopulmonary comorbidities. An age-matched, healthy control cohort was assembled using subjects from the Vascular Model Repository (VMR), an NIH-supported database providing open-access vascular geometries.

### Patient-specific Model Generation

Patient-specific geometries of the pulmonary arterial tree were generated from contrast-enhanced computed tomography (CT) angiography according to previously established methods for the pulmonary vasculature (Figure 1A) [18,21]. SimVascular, an open-source software platform developed for CFD in the vasculature [22] was used to create pathlines across the branching pulmonary arteries. Manual point selection near the vessel center was reviewed by a pulmonologist to ensure pathing that was specific to the pulmonary arteries and excluded veins. Along pathlines, 2D cross-sections were segmented to include the patent lumen of the vessel. In vessels with thrombus, segmentations excluded thrombus from regions of flow. For free thrombus in a given 2D cross-section, an equivalent area was used to represent the degree of occlusion. If a vessel was fully occluded but demonstrated downstream reconstitution of contrast, the vessel was segmented with a minimal connecting channel. Vessel segmentation was halted when a branch met one of two criteria: 1) the vascular lumen was 1 pixel in size or 2) the vessel demonstrated no intralumenal contrast enhancement and did not demonstrate downstream reconstitution in any of its daughter vessels. After segmentation, individual cross-sections were lofted to generate a 3D model. Models were then exported to the open-source mesh-editing software Meshmixer to smooth the intersection of branches, which often contain sharp edges inconsistent with real vasculature. Smoothed models were returned to Simvascular for volume and boundary-layer meshing in TetGen. Mesh-independence of final solutions was confirmed by verifying changes of <5% in pressure and wall shear stress (WSS) for a 15% increase in element number.

**Figure 1.**
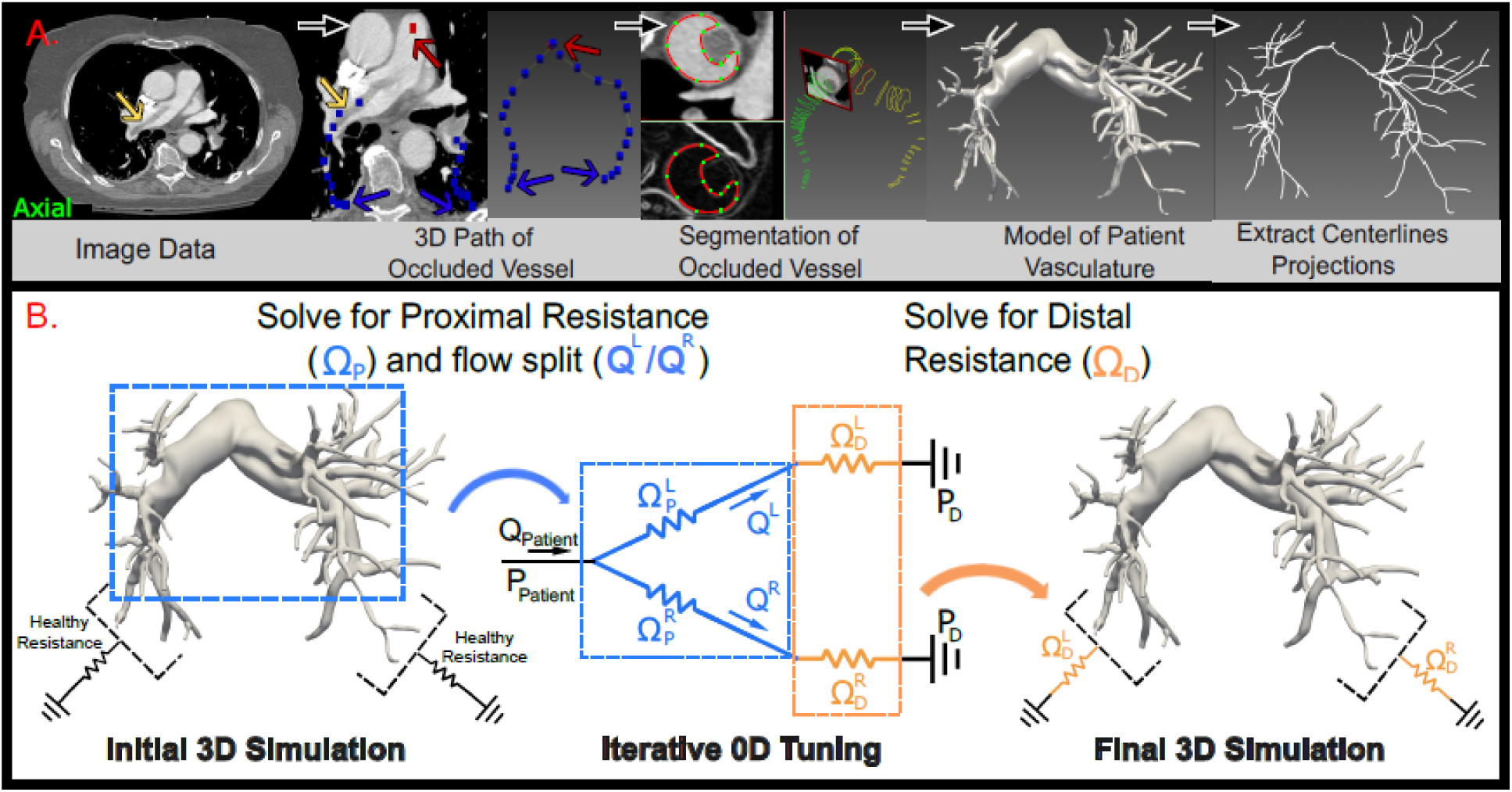
Schematic representation of model building and CFD tuning framework. **A.** Pipeline for generating 3D model for simulation of hemodynamics through importing image data in Simvascular, pathing of patent vessels above the imaging resolution, segmentation of flow path given by contrast, lofting to build water-tight 3D model, and extraction of centerlines for quantification of biomechanical metrics. **B.** Successive boundary condition tuning through initial estimates of 3D (proximal) resistance and flow split, optimization of outflow resistances in reduced order (0D) model, and simulation of 3D flow with tuned boundary conditions giving mPAP matching within 5% of measured clinical target.

### Boundary Condition Tuning

Patient-specific boundary conditions were determined from pre-intervention experimental data for each subject. As there was no flow waveform data, we performed steady simulations with each patient’s CO as the inlet boundary condition. Distal pressure beyond the model boundary was assumed to be 4 mmHg to represent a typical left atrial pressure. Values of resistance for each outlet were determined in an iterative process across multiple tuning steps (Figure 1B). An initial 3D simulation was run using outlet resistances values from the distal vasculature of healthy patients with a typical 55%/45% flow split to the right/left lungs, which were distributed across the outlet faces according to their surface area [23]. Branches with <0.01% of the inlet flow were removed from subsequent iterations. This initial simulation allowed for estimation of the total proximal resistances of the right/left lungs by finding the average pressure drop per flow to each outlet from a given side. These proximal resistances were used in a 0D surrogate model that reduced the contributions of the 3D model to these approximated single resistance values and included single bulk resistances for the distal contribution to each side. Bulk distal resistances were then numerically optimized to match the inlet pressure of the surrogate to the measured mPAP from the clinical data, while maintaining the flow split from the initial simulation with the assumption that proximal lesions primarily drove perfusion. After optimization, a 3D simulation was run with the estimated distal resistances distributed across the 3D model outlets, and deviation from mPAP was examined. If deviation was greater than 5% or 1 mmHg, then this process was repeated with 0D resistances calculated from the updated 3D simulation results, until all deviations fell beneath this threshold.

### Post-processing and Determination of Secondary Metrics

While useful to visualize qualitative differences, hemodynamics across 3D models can be difficult to compare quantitatively. To condense the 3D results onto a simpler geometry for analysis, centerlines from each model were extracted using the VMTK toolbox integrated with Simvascular. Calculation of biomechanical metrics was then performed in Paraview (Kitware, 5.13.1) using its Python scripting interface. Our code for the following centerline projection and metric extraction is publicly available (https://github.com/Carnegie-Mellon-CGRL/Centerline_Programmable_Filters). Centerlines were imported and respaced, dispersing points across 0.01 cm increments while preserving the original geometry. On these centerlines, a number of metrics were derived or projected. A series of programmable filters were used to directly project the pressure and velocity fields from the 3D mesh to the 1D centerlines. Flow was calculated by taking the surface integral of the velocity on the model plane normal to the centerline. This same normal plane was used to find the surface-averaged WSS from the surrounding vessel. In both instances, area of the plane was restricted at junctions to within a range determined by the parent vessel radius.

After values were projected to the centerlines, additional metrics were directly calculated from the reduced values on a segment-by-segment basis, where individual vessel segments were divided at bifurcations. Resistance for each segment was calculated as the total pressure drop from segment inlet to outlet per segmental flow. Segment-wise power dissipation was derived by multiplying the pressure change from the segment’s inlet and outlet by an average flow taken before the segment reached the branching region. A metric related to the commonly used fractional flow reserve (FFR) for the coronary circulation was also calculated as the ratio of the daughter segment terminal pressure to that of its parent vessel.

In addition to these proximal metrics derived from the 3D model, we also examined distal metrics extracted from calculations with the outlet BCs. Specifically, we examined distal resistance as the equivalent resistance to the aggregated left and right BC resistances in a parallel configuration. We also calculated the total distal power dissipation as the sum of the power dissipated by each individual outlet, given by the pressure drop from the 3D model outlet to the terminal pressure multiplied by the outlet flow.

### Statistical Analysis

For the resulting quantified simulation results, three sets of statistical analysis were performed in GraphPad Prism. For HC vs PE comparisons, a two-tailed student’s t-test was performed. To assess differences amongst HC, PPH, and nPPH groups, we used an ordinary one-way ANOVA with multiple comparisons. Finally, to assess correlations between CFD metrics of interest and clinical parameters, we performed a Pearson’s correlation analysis with linear regression. Due to the small sample size of the study, significance was assessed for p<0.1 and all p-values for comparisons are reported in the results section.

## RESULTS

Six patients diagnosed with acute PE and three healthy controls (HC) were included in this analysis. The PE cohort had an age range between 45 and 79 years with comparable HC ages between 40 and 69 years. Pre-treatment transthoracic echocardiography (TTE) data was available for five of six PE patients, all of whom exhibited a dilated right ventricle (RV) with decreased function. McConnell’s sign was positive in four of these patients and negative in one. Invasive pre-operative right heart catheterization of the PE cohort demonstrated hemodynamic impairment prior to intervention (Table 1). Notably, mean pulmonary arterial pressure (mPAP) was >20 mmHg for all patients, ranging from 21 to 34 mmHg. Pre-treatment cardiac index (CI) across the cohort ranged from 1.34 to 2.26 L/min/m^2^. Following treatment, all PE patients demonstrated a clinical hemodynamic response, characterized by a significant absolute reduction in mPAP (ΔmPAP, −9.16 ± 1.08, p < 0.01).

**Table 1.**
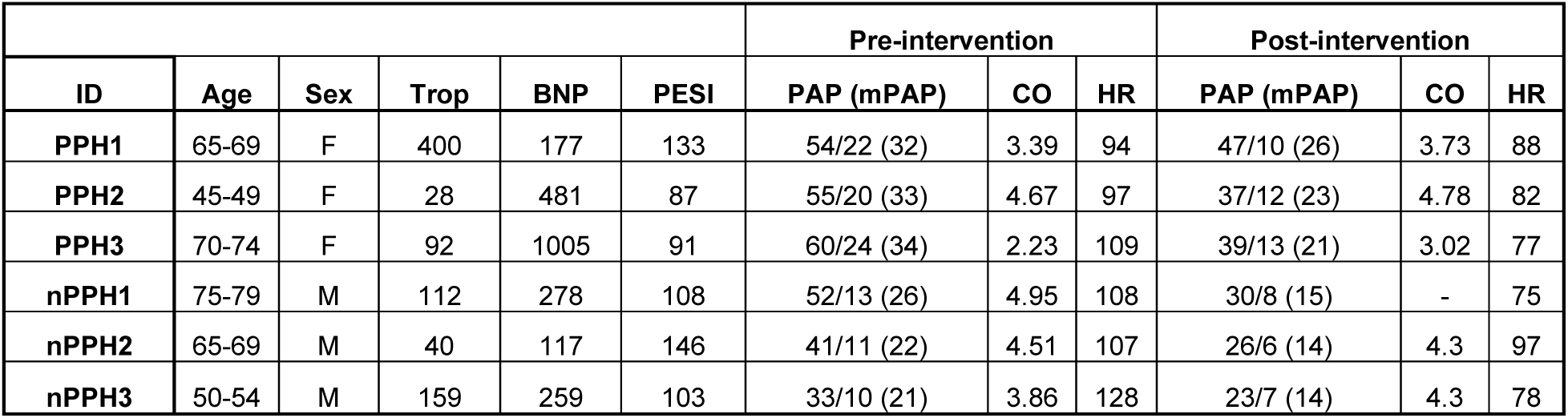
Patient characteristics for the Acute PE cohort. Units for age range (years), troponin (trop: ng/L), B-type natriuretic peptide (BNP: pg/mL), pulmonary embolism severity index (PESI:-) pulmonary artery pressure (PAP: mmHg), cardiac output (CO: L/min), and heart rate (HR: beats/min).

|  |  |  |  |  |  | Pre-intervention |  |  | Post-intervention |  |  |
| --- | --- | --- | --- | --- | --- | --- | --- | --- | --- | --- | --- |
| ID | Age | Sex | Trop | BNP | PESI | PAP (mPAP) | CO | HR | PAP (mPAP) | CO | HR |
| PPH1 | 65-69 | F | 400 | 177 | 133 | 54/22 (32) | 3.39 | 94 | 47/10 (26) | 3.73 | 88 |
| PPH2 | 45-49 | F | 28 | 481 | 87 | 55/20 (33) | 4.67 | 97 | 37/12 (23) | 4.78 | 82 |
| PPH3 | 70-74 | F | 92 | 1005 | 91 | 60/24 (34) | 2.23 | 109 | 39/13 (21) | 3.02 | 77 |
| nPPH1 | 75-79 | M | 112 | 278 | 108 | 52/13 (26) | 4.95 | 108 | 30/8 (15) | - | 75 |
| nPPH2 | 65-69 | M | 40 | 117 | 146 | 41/11 (22) | 4.51 | 107 | 26/6 (14) | 4.3 | 97 |
| nPPH3 | 50-54 | M | 159 | 259 | 103 | 33/10 (21) | 3.86 | 128 | 23/7 (14) | 4.3 | 78 |

Patient-specific 3D CFD models were generated and analyzed to compare the underlying cardiopulmonary biomechanics between the healthy control and PE cohorts. Successful tuning of the resistance BCs gave close matching (<5% error in all cases) of simulated mPAP to the clinical values, which allowed for visualization of the distinct point-wise spatial changes between HC and PE cases (Figure 2). Pressure losses in the branch PAs were generally modest before occlusions that affected the lobar bifurcations (Supplemental Figure 1). In addition to pressure differences, simulations revealed significant elevation of mean WSS across vessel segments (39.2 dyne/cm² vs. 20.7 dyne/cm², p = 0.002) in the Acute PE vs HC cohorts with particularly high values at sites of embolic narrowing (Figure 3). Yet, the presence of wall distortion from an embolus did not always cause elevated WSS levels. In the branch pulmonary arteries, WSS values remained near normal despite clear evidence of clot burden (Supplemental Figure 2).

**Figure 2.**
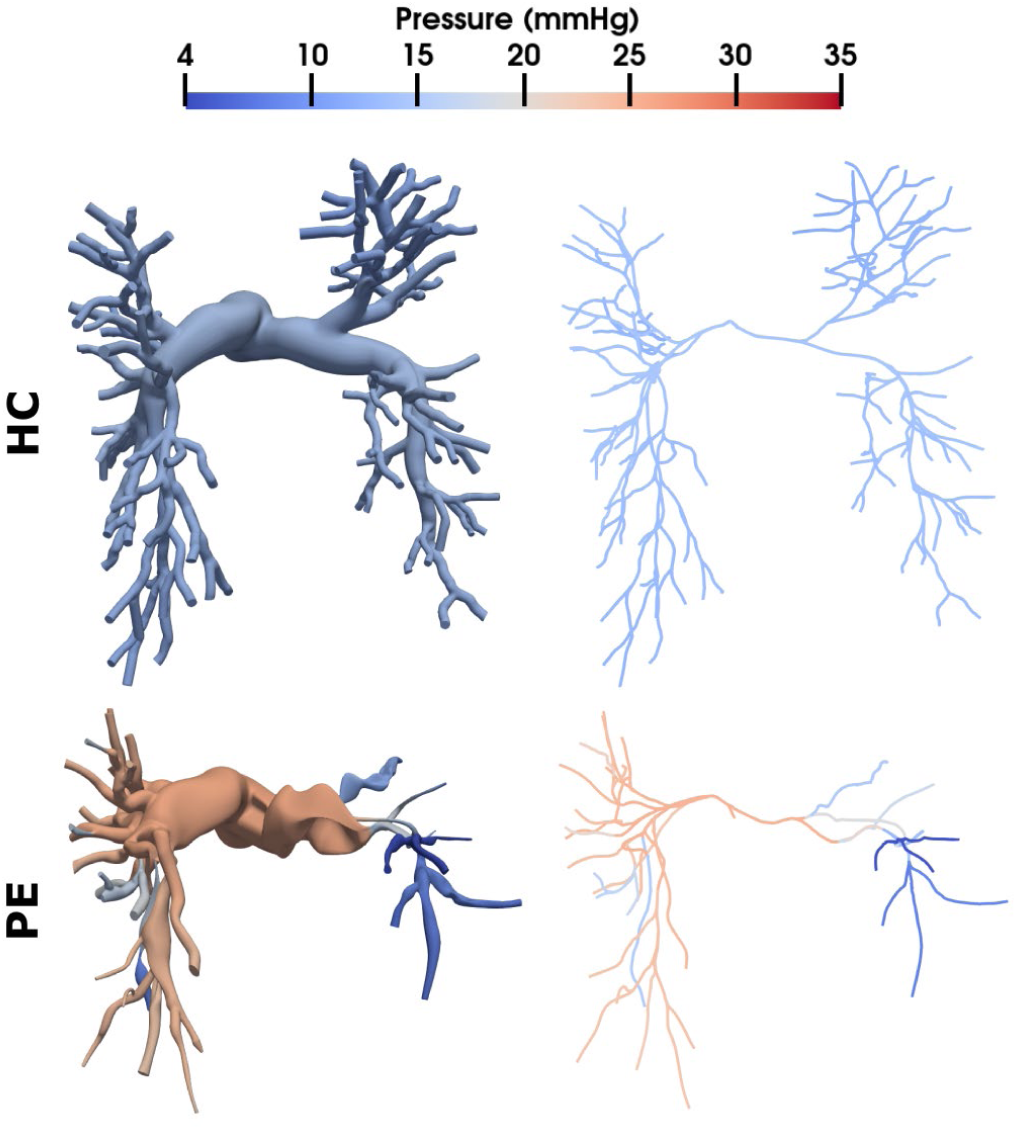
Simulated pressure distribution across HC and PE patients in 3D and centerline models. Pressures were elevated in the PE models with large gradients evident at sites of obstruction in the lobar and segmental PAs.

**Figure 3.**
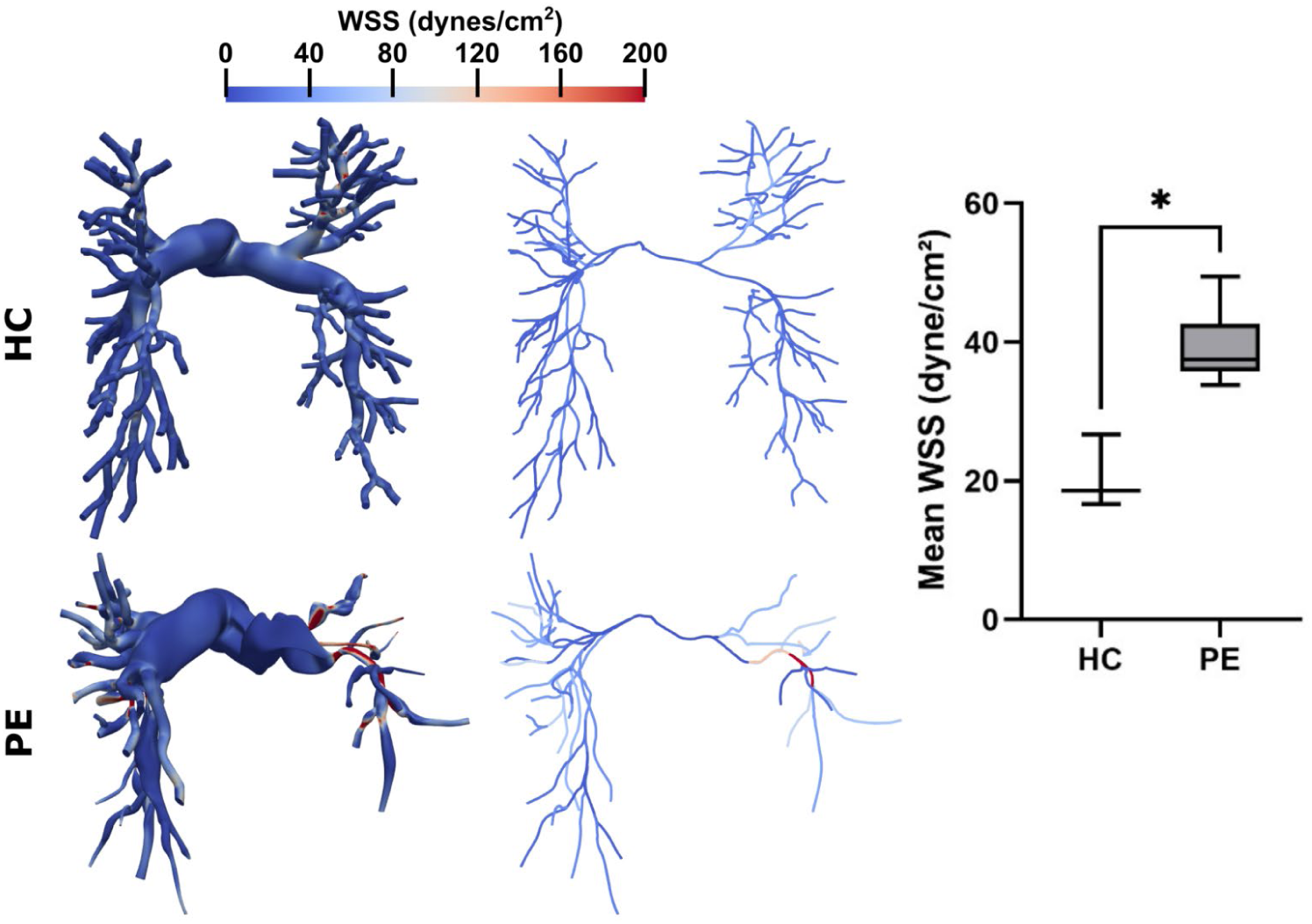
Simulated wall shear stress (WSS) values across HC and PE patients in 3D and centerline models. WSS was lower in the main/branch PAs due to dilatation, but elevated at sites of occlusion. Mean WSS across vessel segments was significantly higher in PE than HCs (p<0.05).

In addition to pointwise estimates, we performed spatial averaging to quantify segment-specific metrics for assessing biomechanical function in the 3D portion of our models. Proximal vascular resistance (144 dyne·s/cm⁵ vs. 20 dyne·s/cm⁵, p = 0.022), total proximal power dissipation (559,814 erg/s vs. 164,905 erg/s, p = 0.010), and the number of low FFR segments (9.67 vs 0, p < 0.001) were all significantly elevated in the PE cohort compared to the HCs (Figure 4). To assess dysfunction in the distal vasculature beyond the imaging resolution of CTA, we examined metrics computed from our optimized boundary conditions (Figure 5). Distal vascular resistance trended higher in the PE cohort, though this difference did not reach statistical significance (361 dyne·s/cm⁵ vs. 134 dyne·s/cm⁵, p = 0.240). In contrast, total distal power dissipation was significantly elevated (1,484,500 erg/s vs. 71,500 erg/s, p = 0.003).

**Figure 4.**
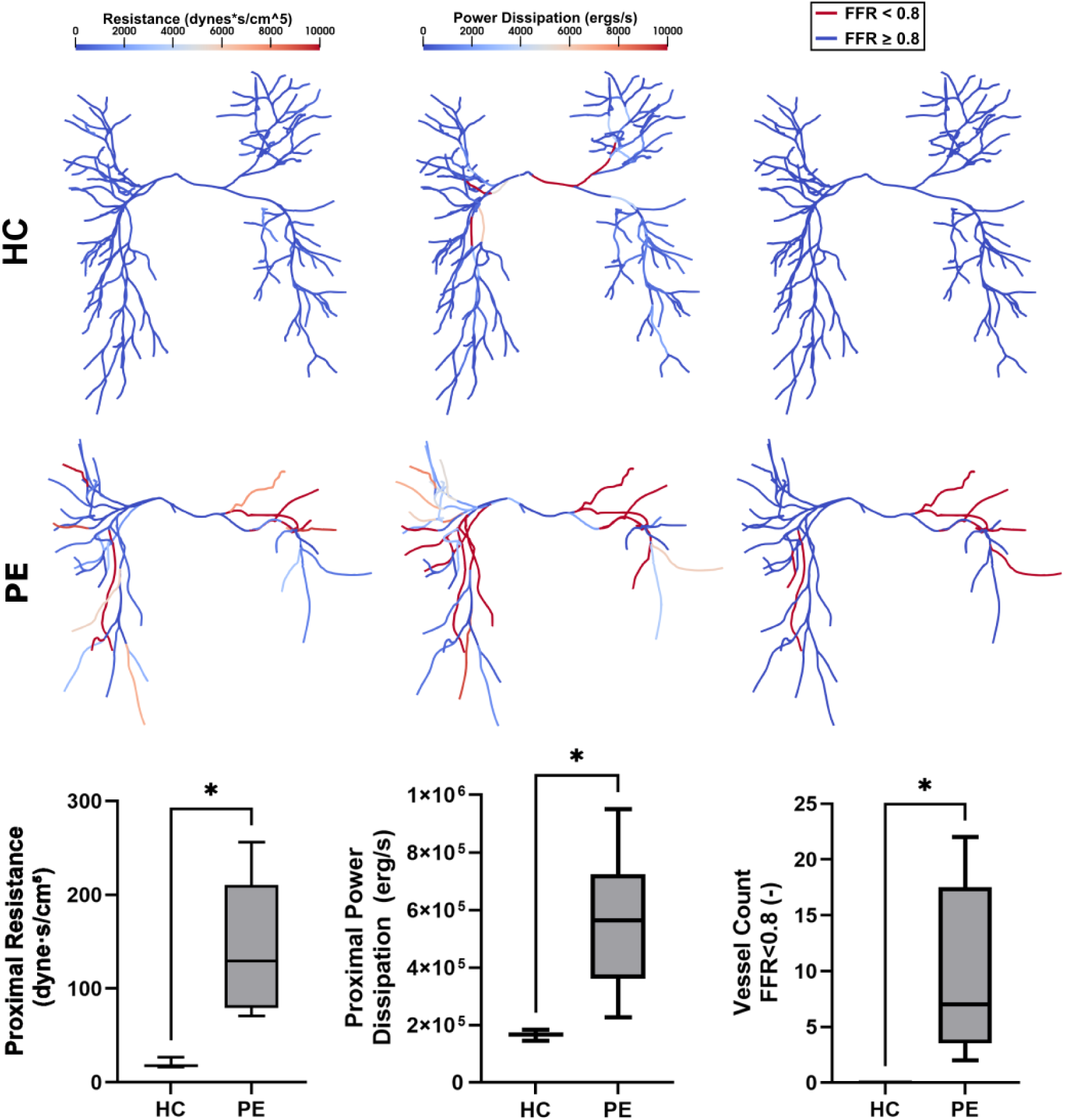
Comparison of proximal metrics of biomechanical function for HC vs PE patients in centerline models. These metrics were derived from the 3D portion of each model. All metrics were significantly elevated in PE patients (p<0.05).

**Figure 5.**
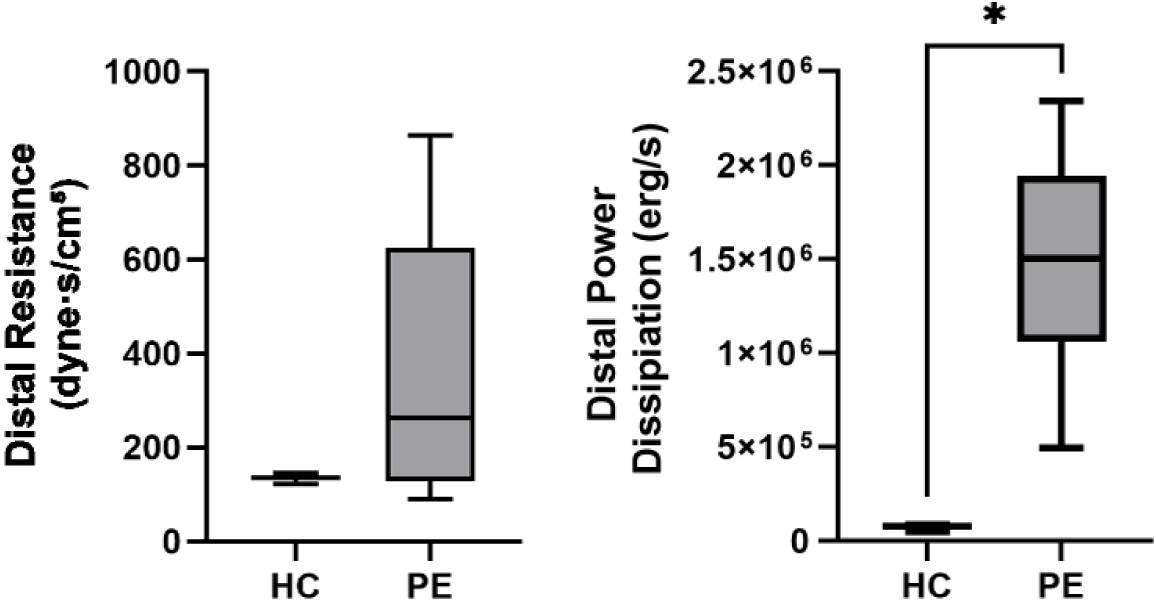
Comparison of distal metrics of biomechanical function for HC vs PE. These metrics were derived from the outflow boundary conditions for each model. Distal power elevation was significantly elevated (p<0.05).

In examining the post-procedural response to thrombectomy, we noted that half of our cohort displayed a residual elevation in mPAP >20 mmHg. Thus, the acute PE cohort was stratified into two distinct subgroups based on post procedural physiology: patients with persistent post-procedural pulmonary hypertension (PPH, n = 3) and patients without post-procedural pulmonary hypertension (nPPH, n = 3). One-way ANOVA revealed significant differences across metrics derived from the segment-based analysis of the 3D models (Figure 6). Proximal resistance was elevated in both diseased cohorts compared to controls, with the nPPH group achieving statistical significance (153.2 vs 20.3 dyne·s/cm⁵, p = 0.075) and the PPH group approaching significance (134.4 vs 20.3 dyne·s/cm⁵, p = 0.121). Proximal power dissipation was elevated in both the PPH (387,543 vs 164,905 erg/s, p = 0.172) and nPPH groups (732,084 vs 164,905 erg/s, p = 0.005) when compared to controls, though the PPH cohort values did not achieve significance in this comparison. Additionally, the nPPH group displayed a significantly higher proximal power dissipation than the PPH group (732,084 vs 387,543, p = 0.056). In examining the number of vessels with FFR<0.8, the nPPH group had significantly higher number of low FFR vessels than the PPH group (16.0 vs 3.67 vessels, p = 0.034) and the HC group (16 vs 0 vessels, p = 0.02) with no difference between the PPH group and the HCs (3.67 vs 0 vessels, p = 0.51).

**Figure 6.**
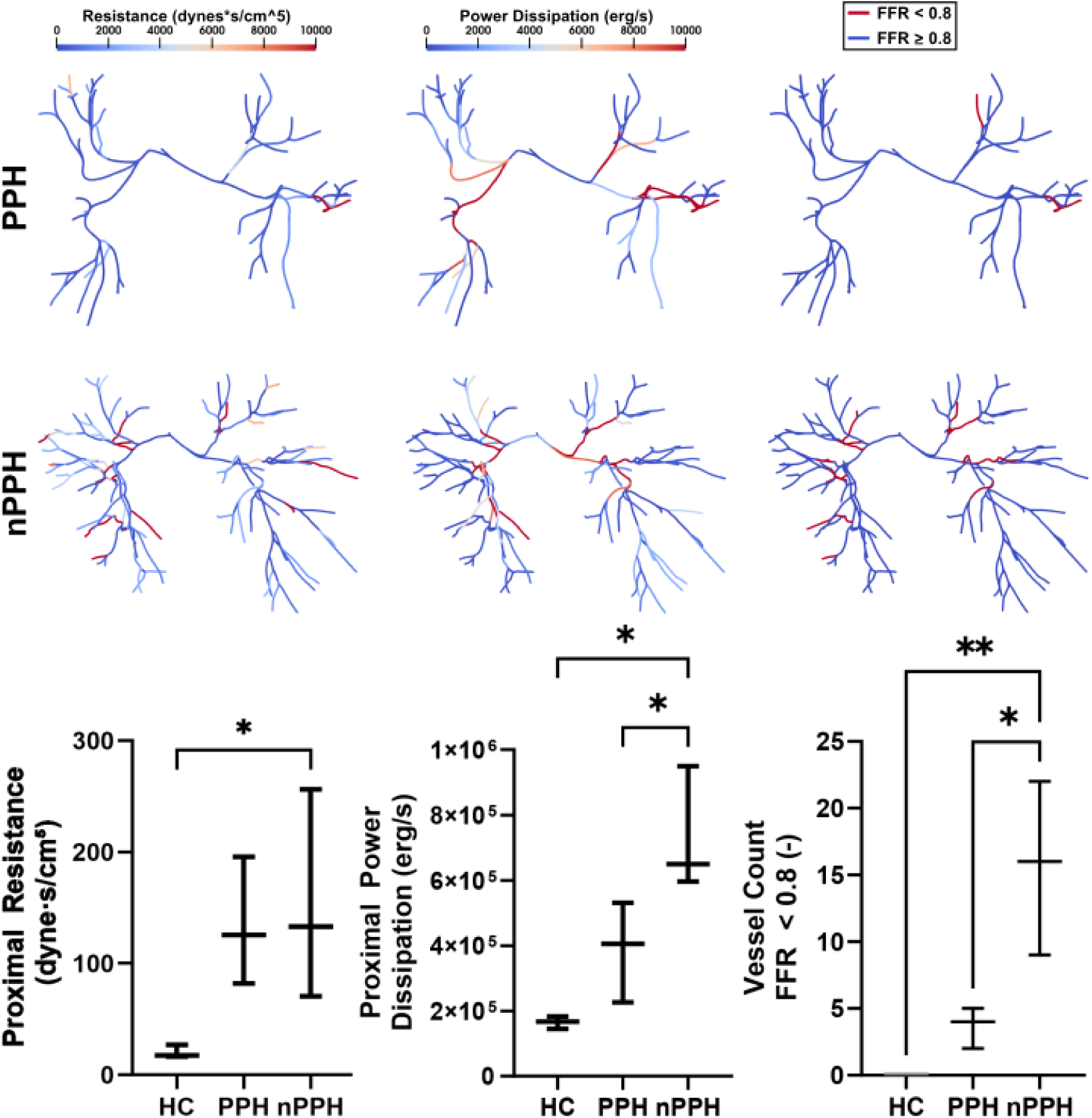
Comparison of proximal metrics of biomechanical function for PPH vs nPPH patients in centerline models. No difference between groups was found for proximal resistance. Proximal power dissipation and number of vessels with FFR<0.8 were both significantly higher in the nPPH group in comparison to the PPH group and HCs (p<0.1).

In addition to examinations of proximal metrics, we compared distal metrics related to our optimized boundary conditions for the nPPH and PPH groups. The PPH group exhibited markedly elevated distal resistance compared to both healthy controls and the nPPH group (583.5 vs 133.9, p = 0.03 and 583.5 vs 139.5, p = 0.03 respectively). No difference was observed for HCs and the nPPH group (139.5 vs 133.9 dyne·s/cm⁵, p = 0.999). Distal power dissipation was significantly elevated in PPH (1,800,000 vs 71,500 erg/s, p = 0.008) and nPPH (1,169,000 vs 71,500 erg/s, p = 0.056) when compared to healthy controls, but no difference was observed between nPPH and PPH groups (1,800,000 vs 1,169,000 erg/s, p = 0.316).

To determine the predictive value of CFD-derived biomarkers, simulated metrics that differed significantly between the nPPH and PPH groups were evaluated against post-mechanical thrombectomy (post-MT) hemodynamic measurements using linear regression analysis, including mPAP and CI (Figure 8). Distal resistance showed a positive correlation with post-MT mPAP (r = 0.69, p = 0.13) and negative correlation with post-MT CI (r = 0.72, p = 0.17), which both approached significance. In contrast, proximal power dissipation demonstrated a negative correlation with post-MT mPAP (r = −0.70, p = 0.12) that approached significance and a significant positive correlation with post-MT CI (r = 0.91, p = 0.03). Finally, the count of vessels with FFR < 0.8 was also negatively correlated with post-MT mPAP (r = −0.9, p = 0.03) and positively correlated with post-MT CI (r = 0.85, p = 0.1) with both metrics showing significance.

## DISCUSSION

There is growing evidence for the efficacy of mechanical thrombectomy for patients with intermediate risk PE, including notable improvements in several outcome measurements in comparison to catheter directed thrombolysis [24]. Yet, after thrombectomy, a significant percentage of patients experience residual symptoms that can lead to longer hospitalization and require chronic monitoring. Additional tools to stratify the risk for persistent post-intervention disease could allow for better intervention planning and targeted therapy. CTA imaging is standard to assess the embolic burden for each patient. There are several quantitative methods to examine embolic burden derived from CTA [25], but their direct clinical utility is mixed. Generally, these metrics do not account for the functional significance of each lesion, i.e. the effect on perfusion and pressure. In this study, we sought to evaluate the use of CFD as a potential tool for characterizing quantitative differences in acute PE and examine its ability to predict hemodynamic outcomes that relate to function.

First, we broadly evaluated the hemodynamic and biomechanical differences between HC and PE groups using patient-specific CFD simulations tuned to match each subject’s clinical data. While inlet pressures and flows were selected to match the clinical data, the evolving pressure and flow distribution through each lesion emerged as a result of the physics-based solution to the governing equations of blood flow (Figure 2). As expected, based on the presence of proximal lesions, the acute PE patients presented with several significant increases in biomechanical metrics related to proximal disease burden. For averaging WSS across vessel segments (Figure 3), the overall value for each patient results from a combination of high-WSS segments with narrowing due to lesions and low-WSS segments that are dilated from increased mPAP. Prior studies have reported global decreases in WSS with embolic pulmonary disease [26], but it is worth distinguishing between averages across segments and the surface average of the whole 3D model used in prior works. Here, we found an increase in mean segmental WSS, which indicates that more individual segments are experiencing increased WSS. Surface averaging of metrics gives outsized weight to the MPA, LPA, and RPA, and obscures the mechanobiological environment at the site of the clot. Increased WSS at each lesion could serve as a stimulus for local endothelial dysfunction and vascular remodeling and contribute to the chronic persistence of lesions [26].

In addition to increased mean WSS, we found increases in proximal vascular resistance, proximal power dissipation, and an elevation in the number of low FFR vessels (FFR<0.8) across the proximal PA tree for acute PE patients in comparison to HCs (Figure 4). These metrics have been used in prior vascular CFD applications, but not in the context of embolic disease. Proximal vascular resistance has been previously used to plan stenting locations in peripheral pulmonary artery stenosis (PPAS) [18]. As in PPAS, narrowing of vessel segments in acute PE patients has a clear physiological implication for elevated resistances. Yet, calculation of proximal resistance is not straightforward, as one cannot only collapse the 3D portion of the model into an equivalent resistive lumped parameter model, but must make an assumption for how it should aggregate. In combining parallel resistances, we found several cases with high unilateral resistances, where the resulting total resistance was then dominated by the vasculature from the lower resistance side of the lung. Power dissipation has not been examined in embolic pulmonary disease previously, though it has been used in congenital heart surgery to minimize the effects of the Fontan conduit in single ventricle patients [27]. In blood vessels arranged in parallel, as in the PAs, it carries a unique advantage relative to resistance in that it can be easily aggregated through summation. Additionally, power dissipation in the vasculature can be directly compared to cardiac power generation for a functional evaluation of how lesions are affecting the workload of the heart. As such, we found increased dissipation of power in proximal vessels of acute PE patients consistent with an increased workload. Finally, examining lesion-specific FFR is widespread in the coronary vasculature as a method for determining the need for angioplasty and stenting in atherosclerosis [28]. CFD-derived FFR from imaging has also allowed for non-invasive characterization of plaque significance and is emerging as a biomarker for severity of lesion narrowing in PPAS and chronic embolic disease for the PAs [28,29]. Here, we extended its use to global evaluation of embolic burden through a lesion count, which revealed an increase in the number of occluded sites in acute PE patients from a total absence of occlusions in HCs.

From our tuned BCs in each patient, we also examined the distal contributions of resistance and power dissipation (Figure 5). Surprisingly, we found no difference in distal resistance between acute PE and HCs, while distal power dissipation was elevated for acute PE patients. On closer examination, there was wider variance in distal resistance than in distal power dissipation across acute PE patients. With subsequent separation of patients according to post-intervention PH status (PPH, mPAP>20 mmHg) vs normotensive post-intervention status (nPPH, mPAP<20 mmHg) (Figure 7), we found a clear elevation in distal resistance only in the PPH group, while those in the nPPH group displayed resistances similar to those in the HC cohort. Distal power across the acute PE groups displayed a smaller difference, though the nPPH group remained lower. This suggested a distinguishing feature between these groups could be the differences in proximal vs distal disease burden. In returning to a comparison of proximal metrics derived from CFD between PPH and nPPH groups (Figure 6), we found that both proximal power dissipation and FFR counts were significantly elevated in the nPPH group compared to the PPH group. Proximal resistances were similar, though the presence of high unilateral resistances in the 3D models could skew results due to the limitations of aggregating 3D resistances noted above. Notably, elevated biomarkers for proximal clot burden was associated with lower post-intervention pressures. This led to a hypothesis that mechanical thrombectomy in intermediate-risk patients is most successful in those patients whose main contribution to disease are in the accessible vessels visible on CTA.

**Figure 7.**
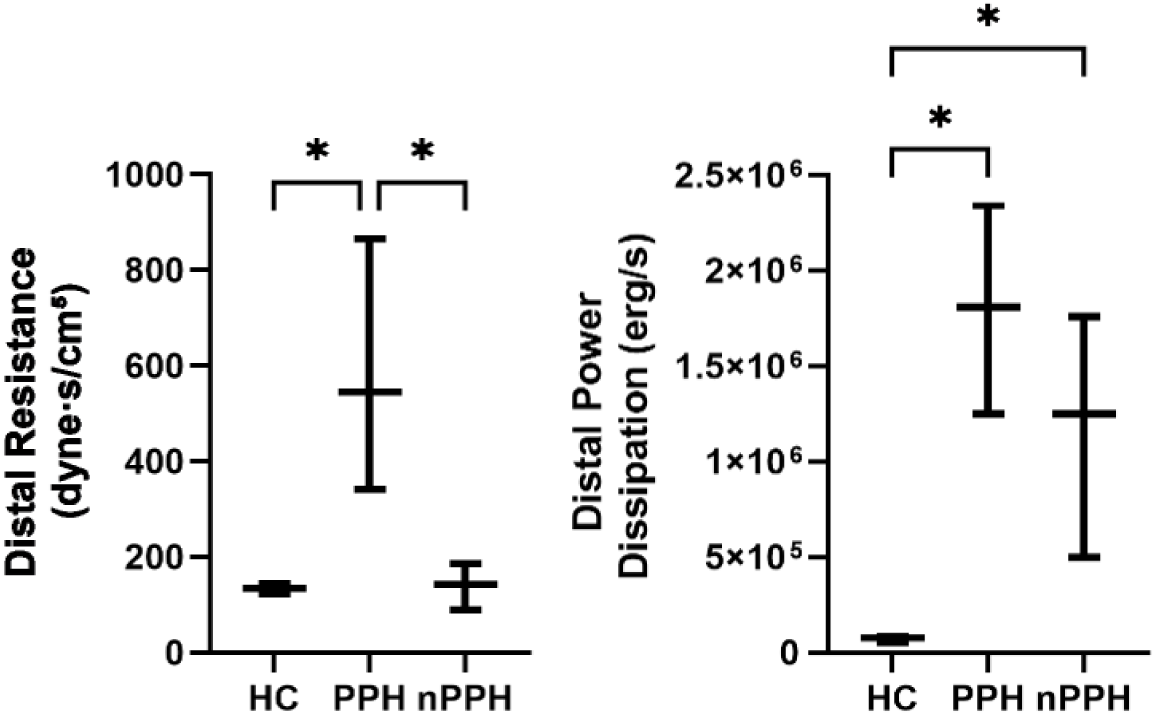
Comparison of distal metrics of biomechanical function for PPH vs nPPH patients. Distal resistance was significantly elevated in the PPH group in comparison to the nPPH group (p<0.05). Distal power dissipation was not significantly different between groups.

**Figure 8.**
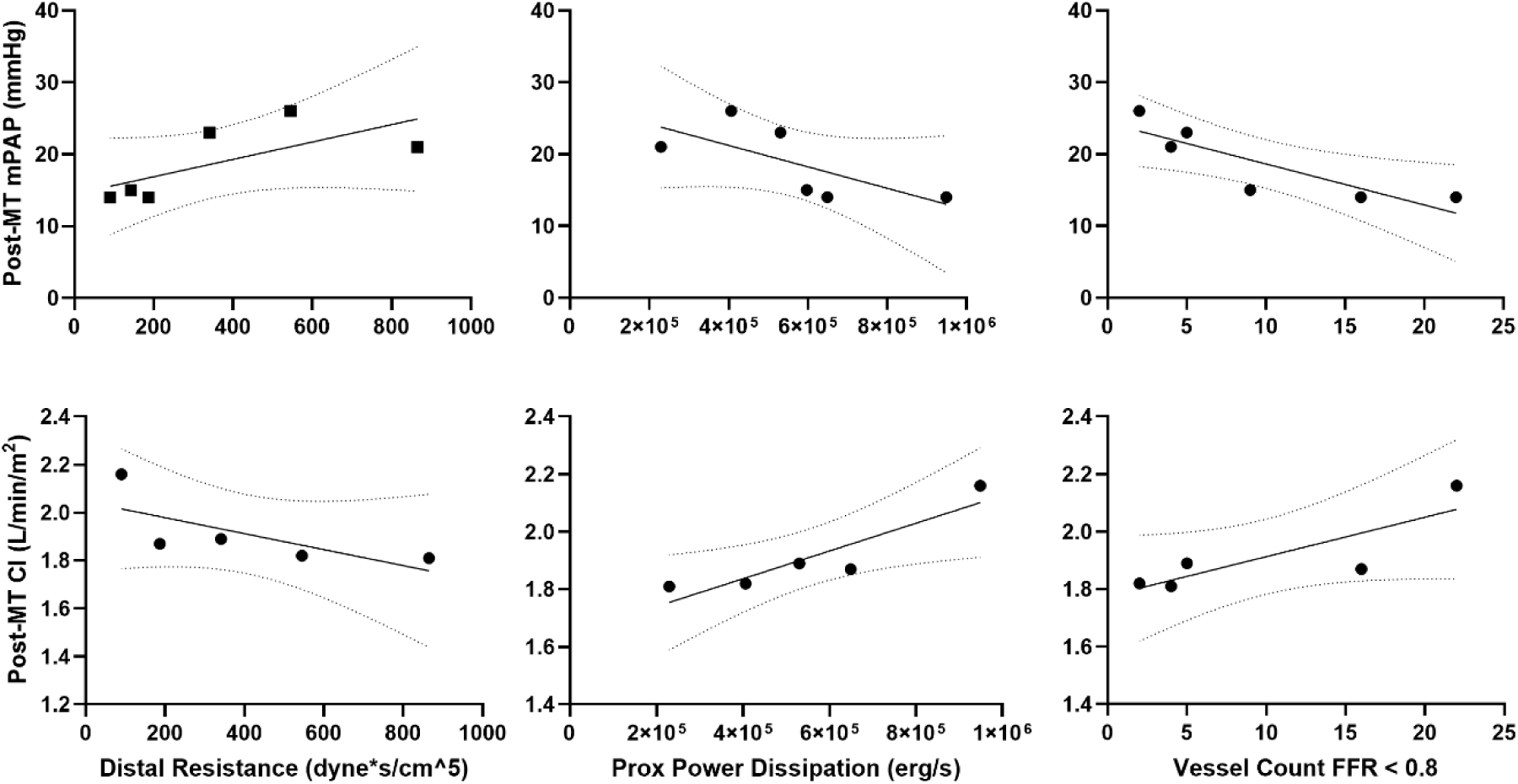
Correlations of significantly different metrics with post-intervention hemodynamics. Proximal power dissipation showed a significant positive correlation with post-mechanical thrombectomy (MT) CI (p<0.05). Vessel count for FFR<0.8 showed a significant negative correlation with post-MT mPAP (p<0.05) and a significant positive correlation with post-MT CI (p<0.1).

In examining the spatial distribution of disease burden through the 3D models, most of the pressure drop and WSS elevations occurred outside of the LPA and RPA despite the presence of large lesions that distorted the luminal cross section (Supplemental Figure 1, Supplemental Figure 2). Instead, these markers tend to be pathologically elevated in emboli located several generations further down the vascular tree. This demonstrates that clot burden in the largest vessels may have a relatively limited contribution to overall clinical severity or acute hemodynamic impairment. Consequently, clot burden at successive downstream generations emerges as a more important marker of disease severity. This requires a reliable way to characterize these target lesions (Figure 9), as proximity to the inlet should not be the sole driver in lesion selection. When evaluating the additional CFD-derived criteria that successfully stratified our subgroup analysis, we found that both FFR and proximal power dissipation varied across PPH and nPPH groups. However, while FFR accurately isolates lesions with a significant impact on local flow, the relative impact on global hemodynamics can remain small if the input flow to that specific vessel is already low, such as in a near total occlusion. Power dissipation, on the other hand, can be erroneously high in areas of shunted, compensatory flow precisely because there is no flow restricting lesion in that region. To address these individual metric shortcomings, we selected for lesions with both high power consumption (in the top quartile of vessel segments for power dissipation) and significant flow restriction (FFR<0.8). These isolated target lesions demonstrated significantly elevated counts in the nPPH cohort compared to the PPH cohort. Furthermore, they tended to be located more proximally within this cohort, as evidenced by the lower median normalized lesion distance in the nPPH group (Figure 9), though the presence of few lesions in the PPH group confounded the statistical assessment of distance. This suggests that further investigation with larger groups may be warranted to examine the effect of embolus location and functional significance on outcomes in mechanical thrombectomy.

**Figure 9.**
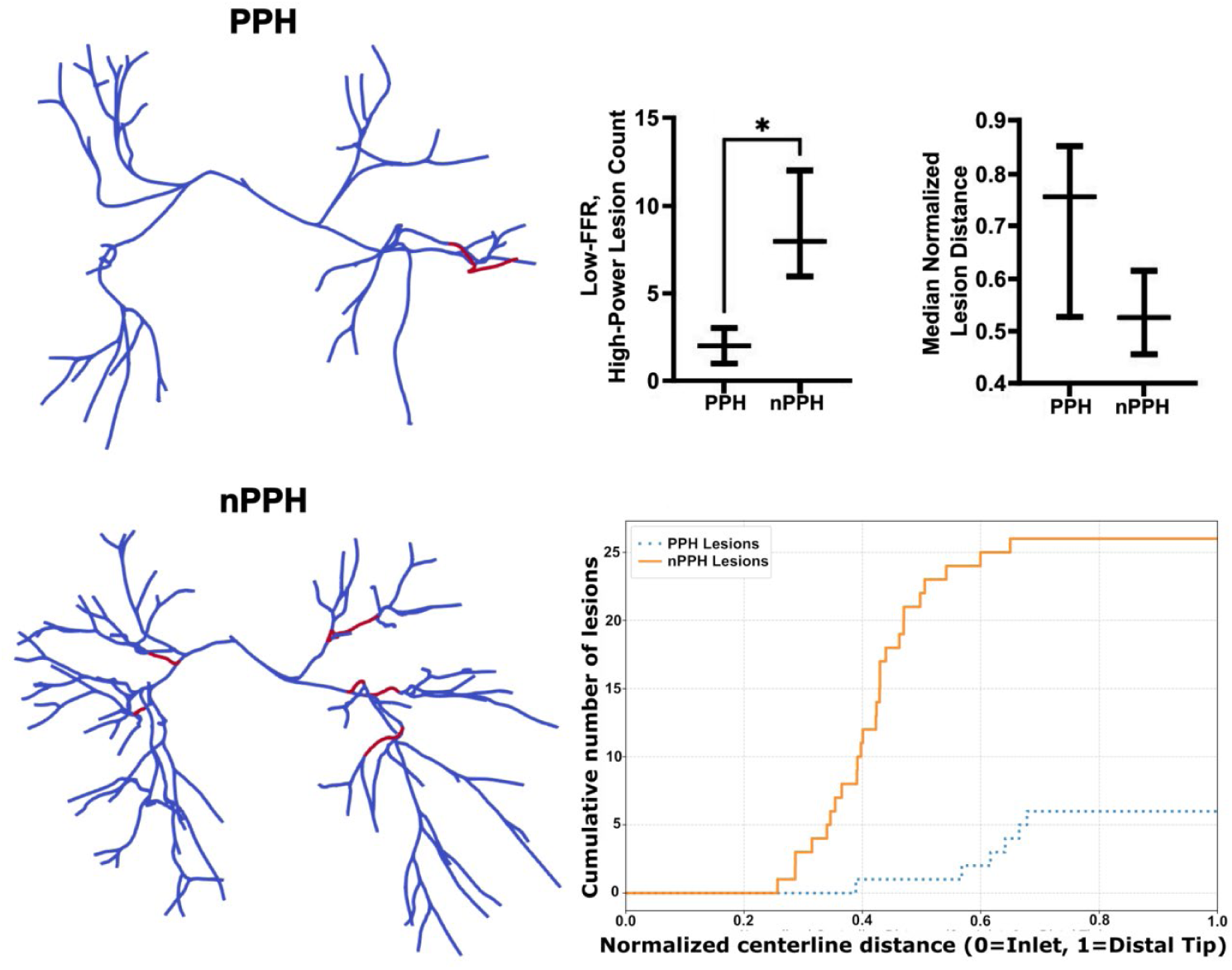
Examination of lesion location on post-intervention status. Number of vessel segments with both low FFR and high power dissipation were significantly higher in the nPPH group (p<0.05), while there was no significant difference in the median distance of lesion across groups. Evaluation of cumulative lesion vs centerline distance for combined groups showed a striking difference in number of lesions across sub-types of PE.

CFD-derived biomarkers have recently been sought to improve predictions of disease development [31] and post-intervention outcomes [32]. As we found significant differences between biomechanical metrics in sub-groups of acute PE patients, we next sought to determine if these metrics could be used as predictors of outcome hemodynamics for future use in stratifying risk for continued dysfunction. Correlations for distal resistance and proximal power dissipation/low FFR count showed opposite trends. This is consistent with the idea that those patients with more proximal disease burden, which is captured by proximal power dissipation and low FFR count, are more responsive to treatment. Those patients with higher power dissipation and higher counts for low FFR vessels tended to have lower post-MT mPAP and higher post-MT CI, suggesting a less diseased state. In contrast, higher distal resistance is unlikely to be affected by MT, leading to more persistent elevation in mPAP and lower CI after MT. Of importance for translation, proximal metrics can be estimated from 3D geometry alone, which offers a path to predicting treatment response in the clinic from imaging prior to catheterization.

Several limitations must be acknowledged in interpreting these findings. First, there was no sex matching; the disease cohort was evenly distributed across males and females, but the healthy control group consisted entirely of females due to the limitations of existing models in the VMR that matched our patient group, and the PPH vs nPPH sub-groups split along female/male sexes. Thus, future studies examining sex-specific differences are necessary. The predictive power and generalizability of these findings are currently limited by the small sample size, necessitating validation in larger, multi-center studies. For the methodology in CFD, the reliance on steady state simulations reduces computational expense but fails to capture the full complexity of pulsatile cardiopulmonary hemodynamics. Indeed, even steady CFD simulations are not useable on a clinical timeline for acute PE, taking several rounds of iterative tuning with runtimes on the order of several days. Additionally, boundary condition tuning relied on maintaining the flow split from the initial simulation with the assumption that proximal lesions primarily drove changes in flow, though the evidence of elevated distal resistances may indicate additional drivers towards flow redistribution. Finally, the absence of exact free thrombus segmentation forces the use of idealized clot geometries such as equivalent areas or minimal connecting channels which may artificially alter local flow separation, wall shear stress, and power loss calculations.

Our study presented several new findings that could guide the use of mechanical thrombectomy and stratify outcomes in acute PE patients. Of key interest, the relative contributions of proximal vs distal disease burden could be used to predict which patients will best respond to intervention. Notably, proximal power dissipation and FFR can be estimated using non-invasive imaging alone, offering a potential pre-intervention tool for planning prior to any catheterization. Furthermore, novel methods for reduced order modeling of the vasculature may offer more rapid simulation times and increase the clinical feasibility of CFD-derived biomarkers to guide real-time patient care. Overall, estimating proximal disease burden from power dissipation or FFR-based biomarkers offers a promising method for prediction.

## Data Availability

All raw data produced in the present study are available upon reasonable request to the authors.

## ACKNOWLEDGMENTS

N/A

## ABBREVIATIONS AND ACRONYMS

BC: Boundary Condition
CFD: Computational Fluid Dynamics
CI: Cardiac Index
CO: Cardiac Output
CPO: Cardiac Power Output
CT: Computed Tomography
CTA: Computed Tomography Angiography
CTEPD: Chronic Thromboembolic Pulmonary Disease
FFR: Fractional Flow Reserve
HC: Healthy Controls
LPA: Left Pulmonary Artery
LV: Left Ventricle
MPA: Main Pulmonary Artery
mPAP: Mean Pulmonary Arterial Pressure
MT: Mechanical Thrombectomy
NIH: National Institutes of Health
nPPH: no Post-Procedural Pulmonary Hypertension
PA: Pulmonary Artery
PE: Pulmonary Embolism
PH: Pulmonary Hypertension
PPAS: Peripheral Pulmonary Artery Stenosis
PPH: Post-Procedural Pulmonary Hypertension
RHC: Right Heart Catheterization
RPA: Right Pulmonary Artery
RV: Right Ventricle
SPECT: Single-Photon Emission Computed Tomography
TTE: Transthoracic Echocardiography
UPMC: University of Pittsburgh Medical Center
VMR: Vascular Model Repository
WSS: Wall Shear Stress

