## Supplemental Figures for "CFD-derived biomarkers in intermediate risk pulmonary embolism patients treated with mechanical thrombectomy"

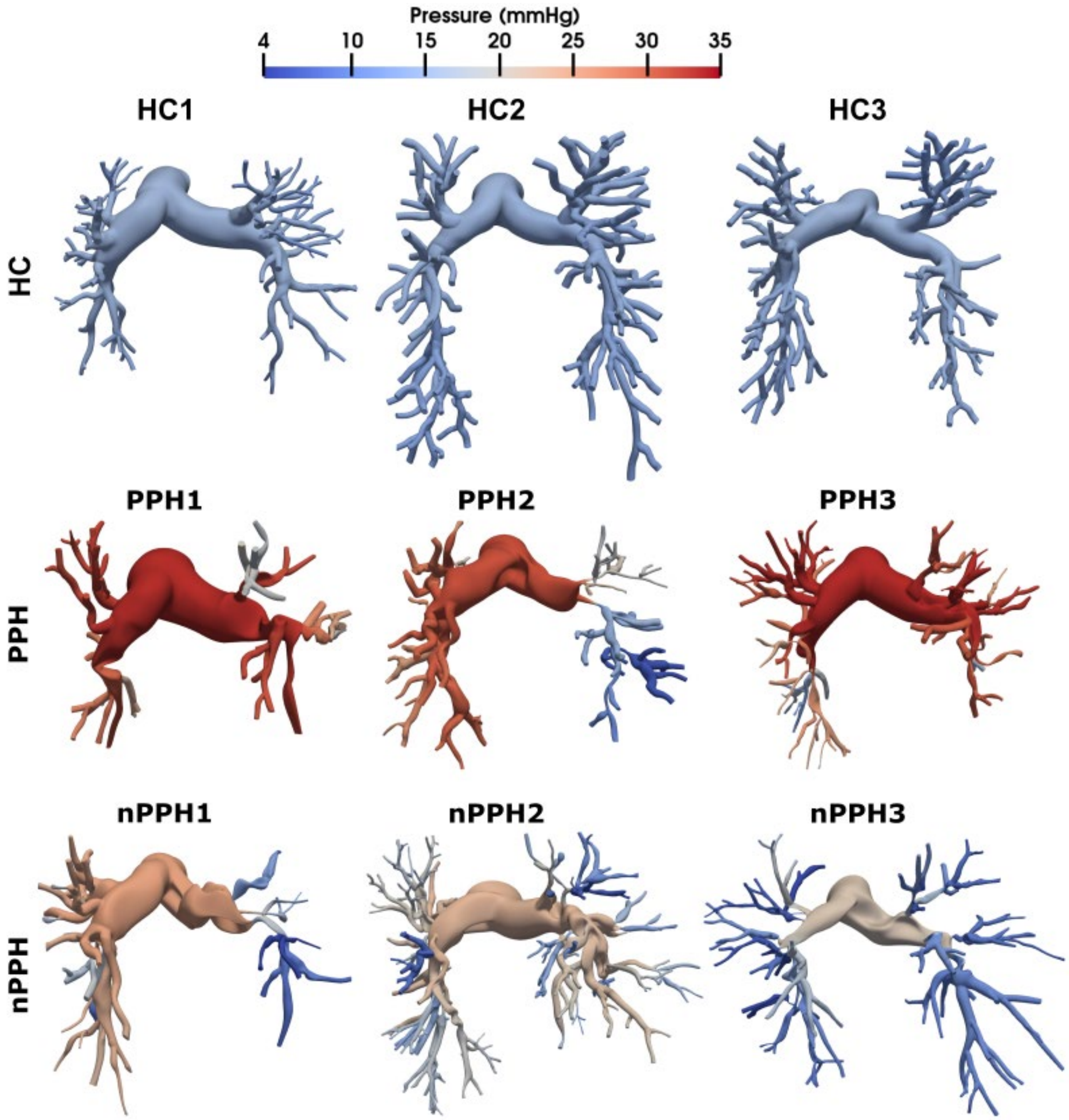


**Supplemental Figure 1.** Pressure distributions for HC patients and acute PE patients, where post-treatment PH (PPH) and no post-treatment PH (nPPH) are separated in the middle and bottom rows. No clear pressure drops were observed in the HC group. For acute PE patients, pressure drops were observed in segmental arteries rather than branch arteries.

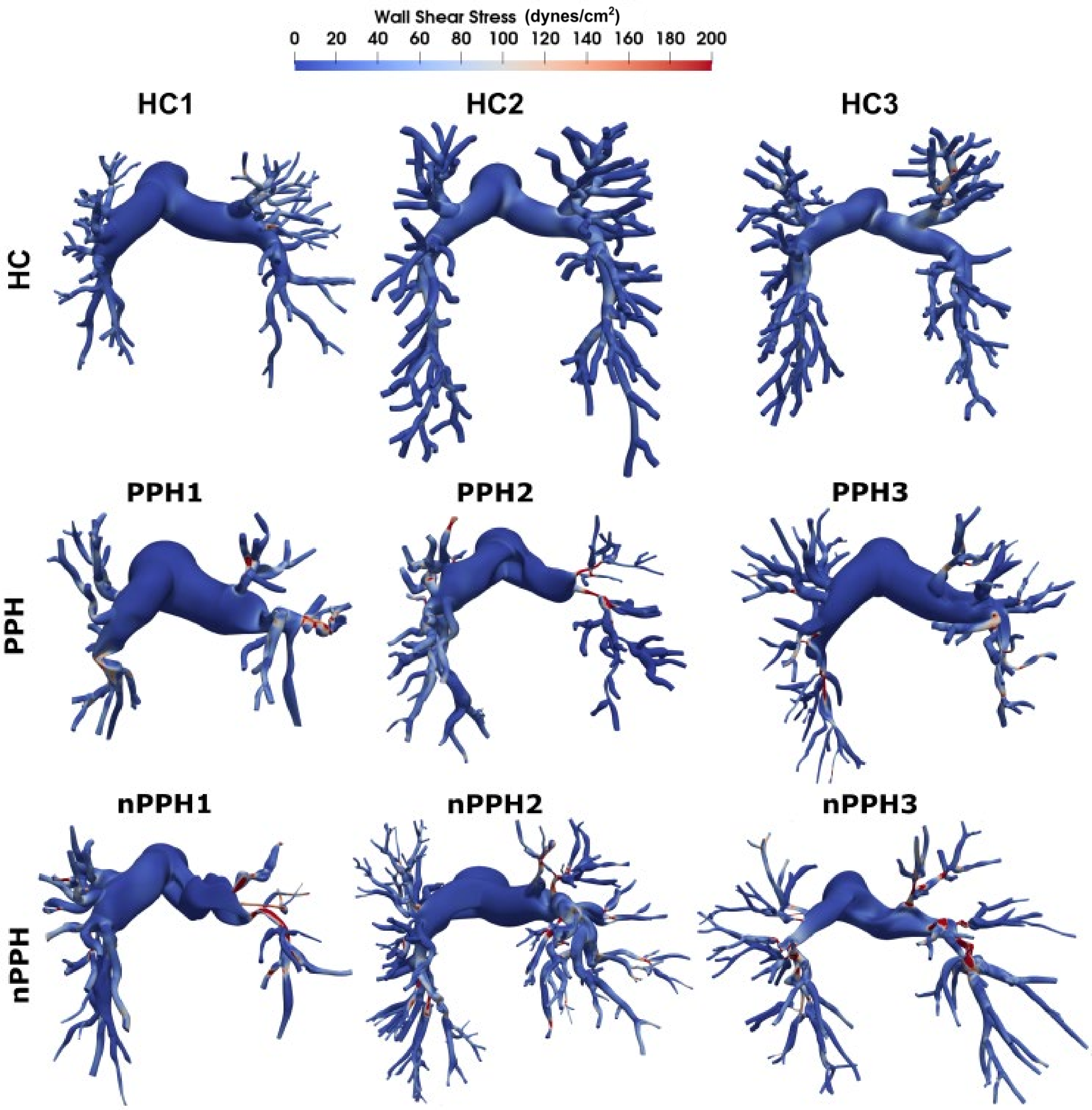


**Supplemental Figure 2.** Wall shear stress (WSS) distributions for HC patients and acute PE patients, where post-treatment PH (PPH) and no post-treatment PH (nPPH) are separated in the middle and bottom rows. Elevated WSS was observed at the site of occlusions in the acute PE cohort, while there were few areas of high WSS in the HCs.
